# Practices and Perceptions Regarding Hyperkalemia in Clinical Practice: A Survey Study in Central America and the Dominican Republic

**DOI:** 10.64898/2026.08.04.26359744

**Authors:** Marta Avellán, Pablo González, Dean Ortíz-López, Josue González, Vicente Sánchez-Polo

**Author notes:** **Corresponding Author**: Dr. Dean Ortíz-López.

## Abstract

**Background:** Hyperkalemia is a clinically relevant disorder across the cardiorenal continuum. In Central America and the Dominican Republic, there are no published systematic descriptions of real-world clinical practices or the degree of alignment of these practices with the most recent hyperkalemia management guidelines.

**Objective:** To characterize physicians’ perceptions and therapeutic behaviors regarding hyperkalemia, including diagnostic thresholds, criteria for intervention and referral, management strategies, and access to potassium monitoring.

**Methods:** A cross-sectional study was conducted using an online survey administered between April and June 2025 to physicians from multiple specialties across seven countries. Absolute and relative frequencies were calculated overall and stratified by specialty and country.

**Results:** A total of 362 responses were collected. Participants were primarily from Costa Rica (32.3%), Honduras (27.9%), and Guatemala (21.0%). 37.8% of respondents reported hyperkalemia in 10%–30% of their patients, with the most reported diagnostic threshold being serum potassium ≥5.5 mEq/L. Outpatient intervention was most frequently initiated at ≥5.5 mEq/L (55.2%), while referral to the emergency department was reported at a potassium level of 6.0 mEq/L (35.6%). Regarding management strategies, 67.0% favored an electrocardiogram prior to deciding on intervention; 93.0% reported reduction or discontinuation of drug causing hiperkalemia; and 74.0% prescribed therapies increasing potassium excretion. Access to potassium monitoring differed substantially by setting, reported as 55.5% in the public versus 90.3% in the private sector. Among cardiologists, frequently used strategies for hyperkalemia in heart failure were reduction or discontinuation of mineralocorticoid receptor antagonists and increased use of loop diuretics. Nephrologists favored strict dietary modifications, loop diuretics, and the use of cation-exchange resins.

**Conclusions:** Substantial heterogeneity was observed in hyperkalemia definitions, action thresholds, and referral criteria, along with frequent modification of renin-angiotensin-aldosterone inhibitors, and reduced access to potassium monitoring in the public sector.

## Introduction

Hyperkalemia (HK) is an electrolyte disorder of high clinical relevance in individuals with chronic kidney disease (CKD) and across the cardiorenal continuum; it is associated with adverse outcomes, increased healthcare utilization, and potentially fatal arrhythmias^1–4^. In clinical practice, HK frequently precipitates dose reduction or discontinuation of therapies with proven cardiorenal benefit—renin–angiotensin–aldosterone system inhibitors (RAASi)—despite their well-established impact on modifying clinical outcomes^1,2^. Persistent heterogeneity exists in the diagnostic and treatment workup of HK, contributing to variability in clinical decision-making ^1,4,5^. The Central American and Caribbean consensus on HK proposes a severity-based categorization (e.g., mild 5.5–5.9 mEq/L; moderate 6.0–6.4 mEq/L; severe ≥6.5 mEq/L) ^6^ along with stepwise management pathways that integrate timely electrocardiographic evaluation, safe acute treatment, and strategies to control serum potassium while reserving RAASi modification as a last resort^1–3,6^. These recommendations are consistent with international guidelines for the evaluation and management of CKD complications, which emphasize monitoring, correction of reversible causes, use of potassium binders and diuretics, and therapeutic continuity if feasible ^7^.

In Central America and the Dominican Republic, real-world clinical decision-making regarding HK—intervention thresholds, management strategies, and approaches to RAASi therapy—has not been systematically described, despite international literature demonstrating substantial variability and gaps between guideline recommendations and clinical practice^1,4,5,8^. This lack of regional data limits local alignment with international and regional consensus documents and may negatively impact on the quality of clinical care delivered to patients. Within this context, the present study aims to determine physicians’ perceptions and therapeutic attitudes toward HK in Central America and the Dominican Republic with the objective of identifying opportunities to improve therapeutic management and education regarding this condition in the region.

## Methods

### Study Design

A cross-sectional study was conducted using a validated and structured regional survey to describe physicians’ perceptions and therapeutic attitudes toward HK in Central America and the Dominican Republic. The survey was developed based on questions previously validated and published in other regions of the world. ^9,10^ The final questionnaire was reviewed and validated by expert representatives from regional cardiology and nephrology scientific societies prior to its implementation.

### Study Population and Inclusion Criteria

The target population included physicians specialized in cardiology, nephrology, internal medicine, endocrinology, geriatrics, and general medicine who were actively engaged in clinical practice across the seven countries of Central America and the Dominican Republic. Physicians contacted for participation were registered in the institutional database of the sponsor and had active informed consent to receive digital communications for scientific research purposes.

### Data Collection Instrument

The 17-item questionnaire was administered using the LimeSurvey platform and consisted of four sections: (1) participant identification, including country and medical specialty; (2) general perceptions, including diagnostic thresholds, use of electrocardiography (ECG), etiological factors, and access to monitoring; (3) therapeutic strategies and decisions regarding RAASi/MRA therapy; and (4) perceived management behaviors in cardiologists for heart failure and in nephrologists for stage 4–5 chronic kidney disease (CKD).

### Procedure

The survey was conducted between April and June 2025. Participants were invited via email, which included a unique link to access the survey platform. Participation was voluntary and anonymous, and access was restricted to a single response per participant. Prior to survey completion, participants were required to confirm their voluntary participation through an electronic informed consent form **(Appendices 1 and 2).** After obtaining voluntary informed consent, the survey was administered. The survey consisted of closed-ended questions, requiring participants to select an answer for each item to progress and complete the questionnaire. To ensure data integrity, only fully completed questionnaires for each of the sections were included in the final analysis.

### Statistical Analysis

Absolute and relative frequencies were calculated for all survey responses. The response rate was defined as the proportion of completed surveys relative to the total number of surveys distributed. Comparisons were carried out across medical specialties and countries. Statistical analyses were conducted using R software 4.0.2 (R Foundation for Statistical Computing, Vienna, Austria).

## Results

The survey was filled by 362 physicians, with only 3 physicians (1 cardiologist, 2 nephrologists) that provided incomplete surveys and are only included in the first section of the analysis. The countries with the highest proportion of respondents were Costa Rica (32.3%; 117/362), Honduras (27.9%; 101/362), and Guatemala (21.0%; 76/362). The most frequently represented medical specialties were internal medicine (27.1%; 98/362), nephrologist (24.3%; 88/362) and general medicine (21.5%; 78/362) (**Figure 1**).

**Figure 1.**
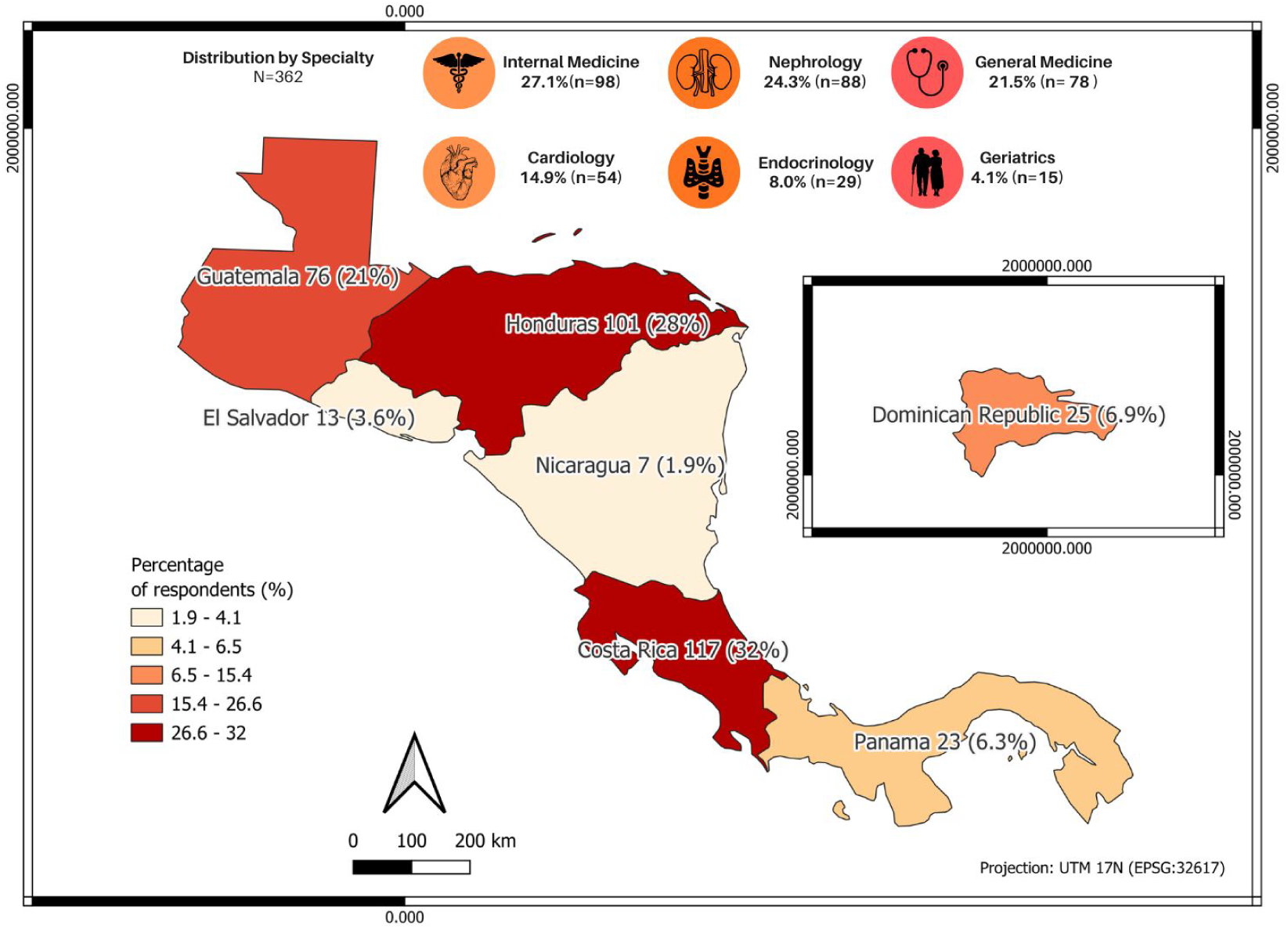
Distribution of surveyed physicians by country and medical specialty.

### Frequency of HK in Clinical Practice

In response to the question, *‘According to your clinical experience, how frequent is hyperkalemia among the patients you routinely manage?’*, most participants (137; 37.8%) selected the option *‘Between 10% and 30% of my patients.’* The second most frequent response was *‘Less than 10% of my patients,’* by 107 respondents (29.6%). This pattern was consistent across all professional groups surveyed, including cardiologists’ perceptions among patients with heart failure. In nephrology, the two most frequently selected responses were *‘Between 10% and 30% of my patients’* (33,37.5%), followed by *‘Between 30% and 50% of my patients’* (29,33.0%) (**Figure 2**).

**Figure 2.**
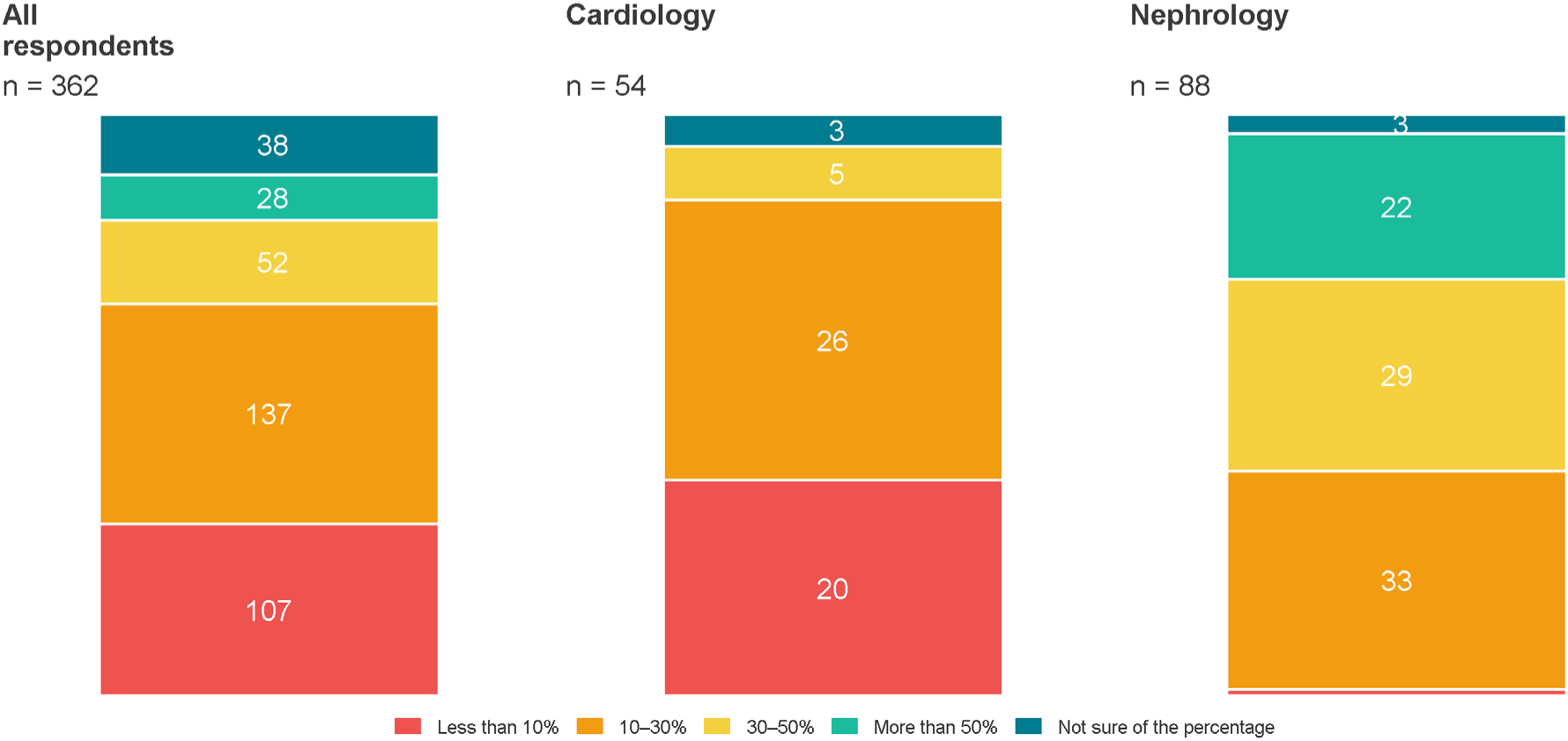
Surveyed physicians according to the perceived proportion of patients with HK in clinical practice.

### Serum Potassium Levels for the Diagnosis and Management of HK

The serum potassium level most frequently reported as diagnostic for HK was 5.5 mEq/L (overall 61.9%; cardiology 57.4%; nephrology 65.9%). Endocrinology was the only specialty with a different pattern, with 48.3% (14/29) of respondents reporting 5.0 mEq/L as the threshold. Regarding the serum potassium at which outpatient medical intervention was considered necessary, the most frequently reported cut-off was 5.5 mEq/L, selected by 55.2% (200/362) of respondents, followed by 5.0 mEq/L in 17.1% (62/362) (**Figure 3, Supplementary Table 1**).

**Figure 3.**
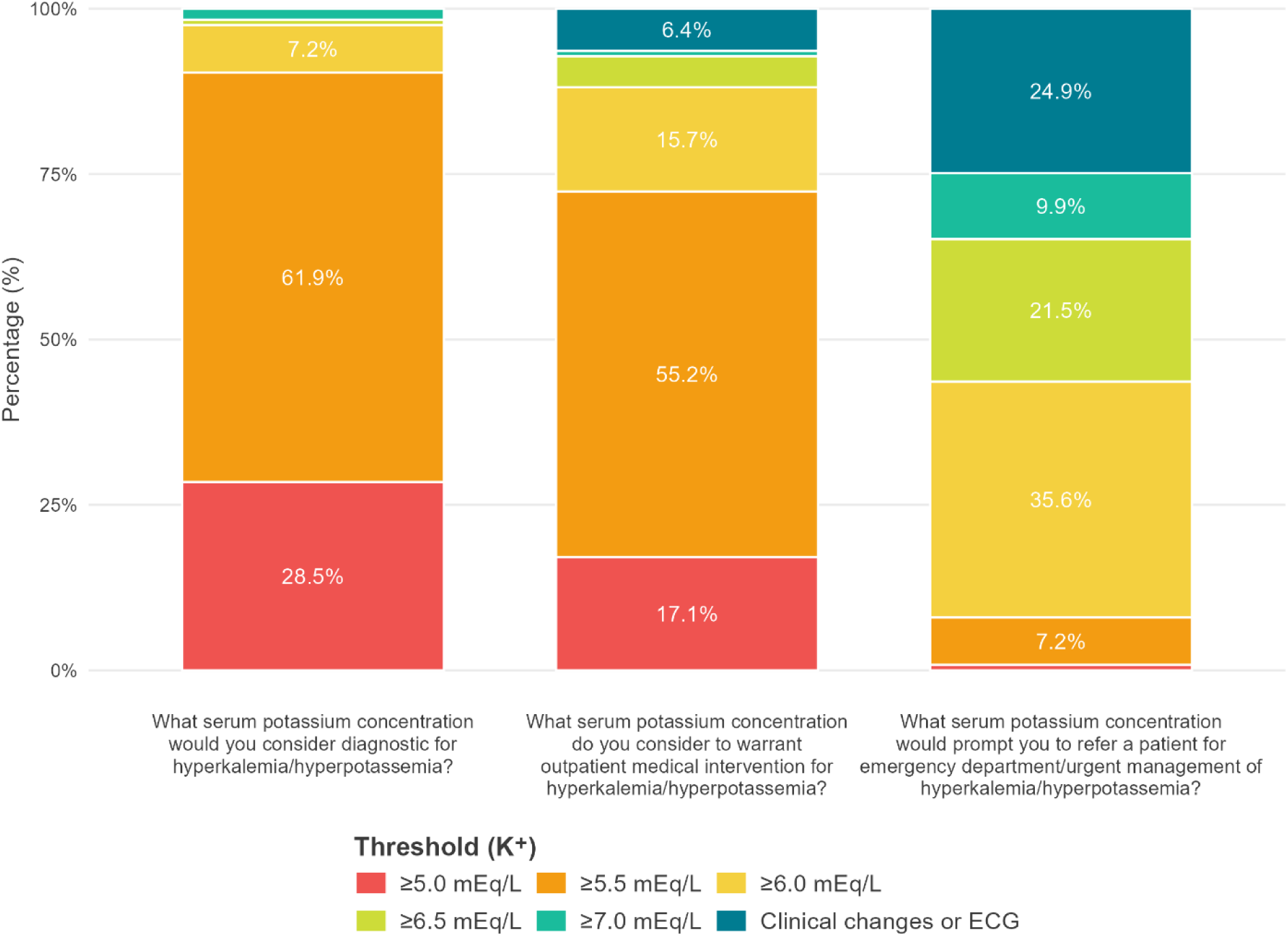
Distribution of physician responses regarding serum potassium levels for diagnosis, outpatient management, and emergency referral for HK.

With respect to the serum potassium level prompting referral to the emergency department for HK management, a value of 6.0 mEq/L was most frequently reported, accounting for 35.6% (129/362) of responses. This was followed by referral based on the presence of clinical or electrocardiographic changes regardless of serum potassium level (90; 24.9%). Both cardiologists and nephrologists most frequently selected the 6.0 mEq/L threshold (44.4% and 33.0%, respectively) **(Figure 3**, **Supplementary Table 1).**

### Perceived Causes and Therapeutic Management Strategies for HK

The option perceived to be a *‘very frequent’*cause of HK by most of physicians was chronic kidney disease (72.7%) while RAAS inhibitors and untreated diabetes were classified as ‘*moderately frequent’* (36.7% and 31.2% respectively) **(Supplementary Table 2)** When asked, *‘Is it routine in your practice to perform an electrocardiogram before deciding whether your patient requires an intervention for elevated serum potassium?’*, 67.0% (241/362) of respondents selected *‘Yes.’* This response was homogeneous across most specialties, except for geriatric medicine specialists, in which only 33.3% (5/15) selected *‘Yes’*. Regarding therapeutic management, reducing or discontinuing medications potentially contributing to elevated serum potassium was selected by 93.0% (336/362) respondents. Among them, 39.5% reported directly discontinuing medications, while 53.3% preferred dose reduction. **(Figure 4, Supplementary Table 3**).

**Figure 4.**
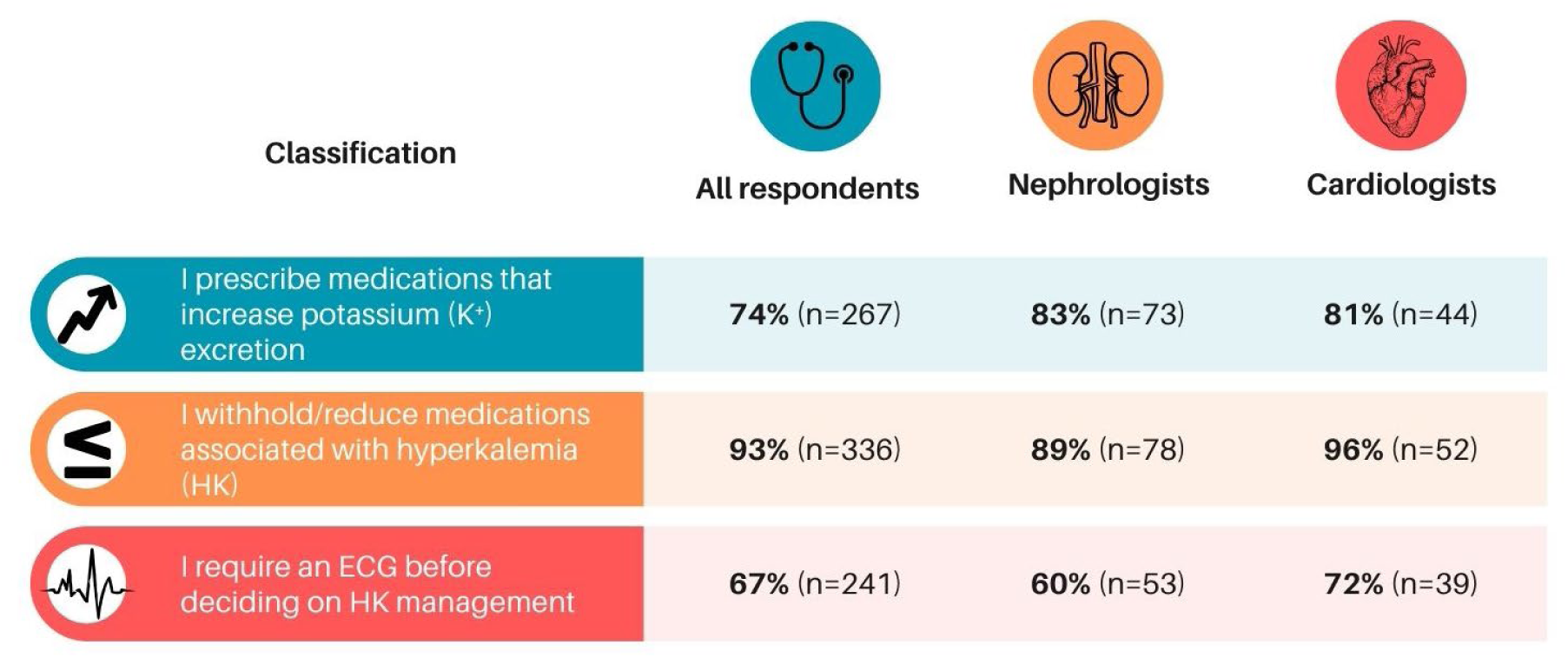
Distribution of surveyed physicians according to HK management strategies by medical specialty.

The specialties most frequently opting for medication discontinuation were internal medicine (57.1%; 56/98), followed by cardiology (42.6%; 23/54). Preference for dose reduction without discontinuation was highest in geriatrics (73.3%; 11/15) and endocrinology (62.1%; 18/29). Another therapeutic strategy assessed was the prescription of medications that increase potassium excretion (e.g., loop diuretics, thiazide diuretics, cation-exchange resins), which was reported by 74.0% (267/362) of respondents. This practice was most frequent among nephrologists (83.0%; 73/88) and cardiologists (81.5%; 44/54). (**Figure 4, Supplementary Table 3**).

### Access to Potassium Monitoring in Public and Private Healthcare Settings

In response to the question, *‘Based on your current practice conditions, do you consider that your patients have access to potassium monitoring as recommended by clinical practice guidelines in the public and private settings?’*, lack of access in the public sector was reported by 44.5% (161/362) of respondents, compared with 9.7% (35/362) in the private sector. Costa Rica and the Dominican Republic reported the highest levels of access to potassium monitoring in the public healthcare system (73.5% and 72.0%, respectively). In contrast, other countries reported similar proportions of access and lack of access (Guatemala, El Salvador) or predominantly lack of access (Honduras, Nicaragua, Panama). The groups reporting lack of access most frequently were general practitioners (57.7%) and internists (49.0%) (**Figure 5, Supplementary Table 4**).

**Figure 5.**
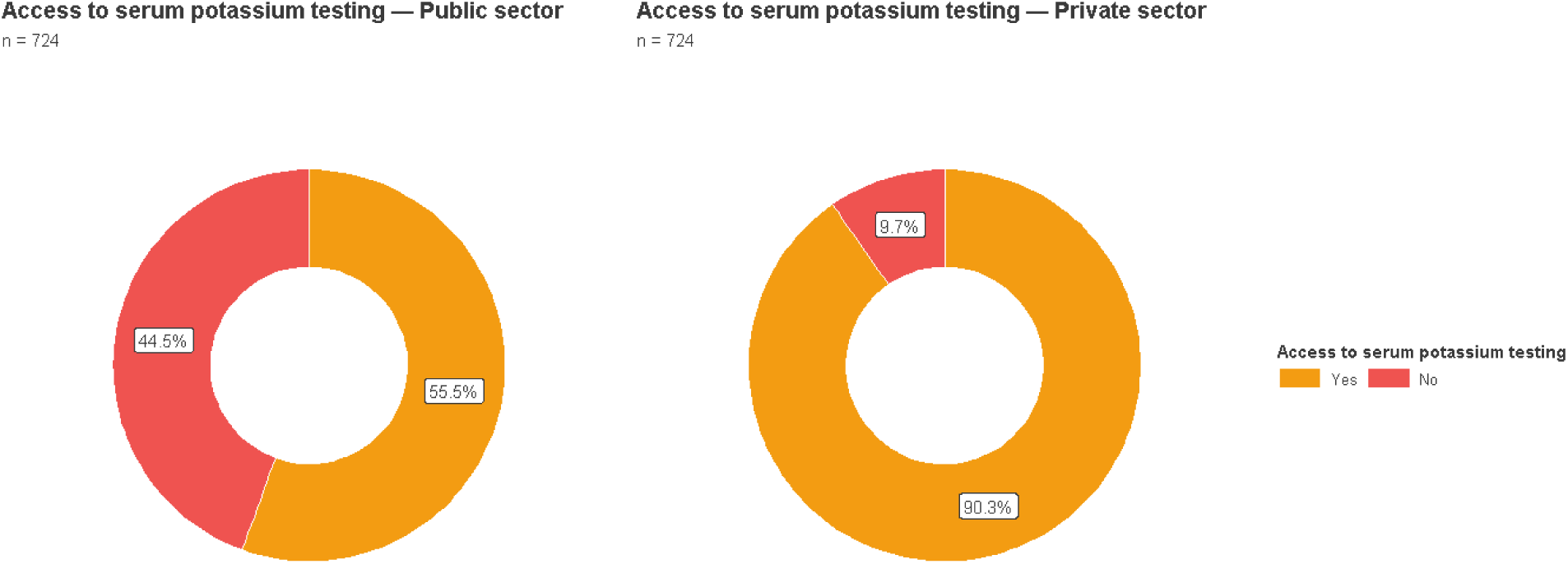
Perceived accessibility to potassium monitoring in public and private sector.

### Cardiology Section: Management of HK in Heart Failure

Among the cardiologists surveyed, 58.5% (31/53) rated HK as a major concern in heart failure, while 41.5% (22/53) rated it as a secondary concern. Regarding pharmacological management strategies, the response *‘Yes, I use it frequently’* was most selected for two options: reduction or discontinuation of mineralocorticoid receptor antagonists (40/53; 75.5%) and initiation of loop diuretics (26/53; 49.1%). The response *‘I use it, but only in some cases’* was most frequently selected for reduction or discontinuation of ACE inhibitors/ARBs (62.3%) and angiotensin receptor–neprilysin inhibitors (60.4%). With respect to cation-exchange resins, 79.2% of respondents reported either not using them or using them only in selected cases (39.6% for each option) (**Figure 6, Supp. Table 5**).

**Figure 6.**
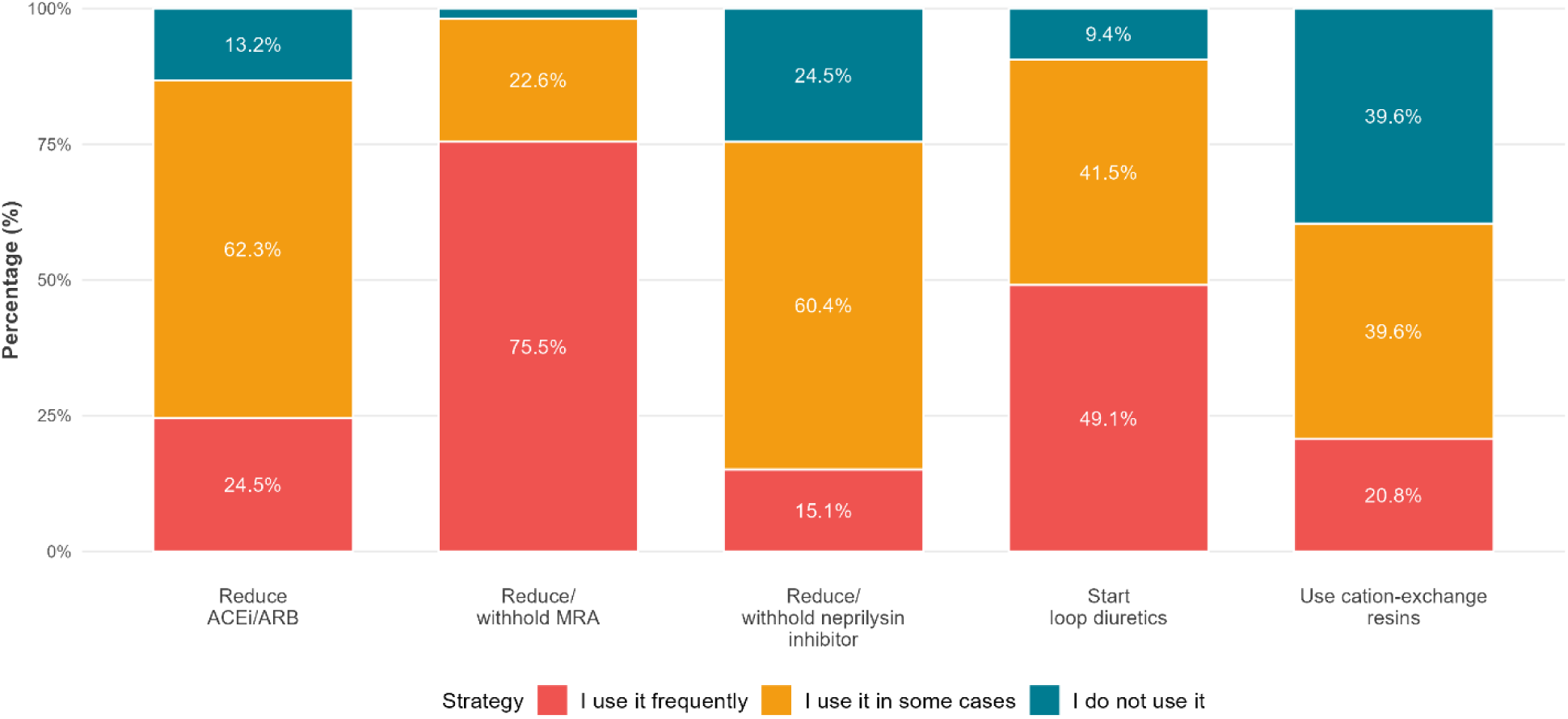
Distribution of cardiologists according to pharmacological management of HK.

### Nephrology Section: HK in Stage 4–5 Chronic Kidney Disease Not on Dialysis

Among nephrologists, the response *‘Yes, I use it frequently’* was most selected for four strategies: use of loop diuretics (67.4%; 58/86), use of cation-exchange resins (62.8%; 54/86), strict dietary restriction (80.2%; 69/86), and observation with repeat monitoring (77.9%; 67/86). The response *‘I use it, but only in some cases’* was most frequently selected for reduction of mineralocorticoid receptor antagonists and ACE inhibitors/ARBs (53.5%; 46/86 and 57.0%; 49/86, respectively) (**Figure 7**).

**Figure 7.**
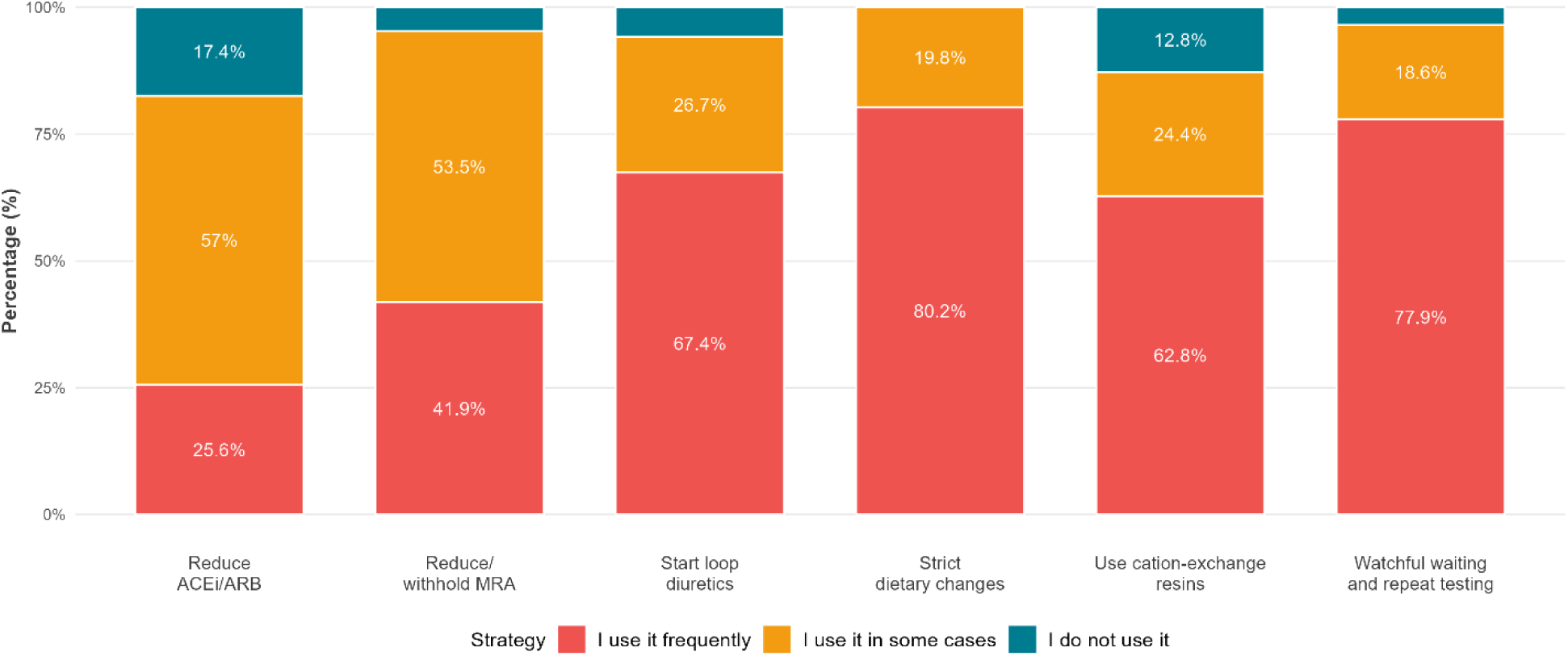
Distribution of surveyed nephrology specialists according to pharmacological management of HK.

## Discussion

In this regional survey, we collected information from 362 healthcare providers from multiple specialties involved in the diagnosis and management of HK. Regarding diagnostic thresholds of HK, over half of respondents agreed on a serum potassium value >5.5 mEq/L, consistent with the Central American Consensus on HK as well as other regional and international consensus documents ^6,9,10^. It is noteworthy that 38.1% of surveyed physicians reported different criteria, particularly among cardiologists, where one in three respondents reported a threshold of 5.0 mEq/L, possibly reflecting adherence to the HK recommendations published by the European Society of Cardiology in 2018 ^10^.

An international survey conducted among 500 primary care physicians also reported 5.0 mEq/L as the most frequently selected threshold ^10^. These findings are directly related to the lack of an internationally standardized definition of HK, a limitation acknowledged by KDIGO, which recognizes that a clinically meaningful definition should be based on the population distribution of serum potassium levels. Epidemiological studies suggest that adverse clinical outcomes— including hospitalizations, all-cause mortality, and decline in kidney function—increase in frequency starting at serum potassium levels of 5.0 mEq/L compared with normokalemic controls ^11–13^. Collins et al., for example, described that for every 0.1 mEq/L deviation outside the range of 4.0–5.0 mEq/L, serum potassium levels were associated with an increased risk of all-cause mortality. These data underscore the need for clearer definitions of HK that integrate both serum potassium values and their clinical implications.

With respect to serum potassium thresholds prompting emergency management of HK, greater heterogeneity in responses was observed. The most frequently selected options were referral at potassium levels ≥6.0 mEq/L (35.6%), ≥6.5 mEq/L (21.5%), or referral only in the presence of electrocardiographic changes (24.9%). In this instance, the Central American Consensus, the KDIGO guideline and the UK Renal Association provide clear guidance on emergency management of HK: for potassium levels ≥6.0 mEq/L, hospital evaluation and treatment are recommended in unstable patients or in the presence of acute kidney injury, whereas stable patients should undergo dietary and medication review with repeat testing within 24 hours^14^. These recommendations are supported by clinical studies demonstrating that patients receiving emergency care with potassium levels >6.0 and >6.5 mEq/L are associated with a three- and six-fold increase in mortality risk, respectively^15^. The results of this survey highlight a lack of clarity regarding which HK-related criteria should prompt emergency referral, a critical issue in a region where access to high-quality emergency care remains heterogeneous.

Another key finding of this study was the high frequency of medication reduction or discontinuation reported by more than 90% of respondents. This approach, although recommended by guidelines as an initial step in HK management, is specifically for medications other than RAAS inhibitors. Nevertheless, the cardiology and nephrology-specific sections of the survey revealed that reduction or discontinuation of RAASi therapy was favored over alternative strategies. This pattern mirrors findings from a previous international survey among cardiologists regarding heart failure treatment, in which approximately 72% of respondents reported modifying RAASi therapy as the primary strategy to manage HK, while fewer than 14% reported using alternative approaches^15^. Modification of RAASi therapy has been associated with increased mortality and hospitalization rates in patients with heart failure, chronic kidney disease, and diabetes in prior studies ^16,17^. More recently, the use of cation-exchange resins has been promoted as an alternative approach to control HK while optimizing RAASi therapy, although barriers such as tolerability, adverse effects, and cost persist. These findings emphasize the need to disseminate alternative strategies to RAASi reduction for HK management to avoid unnecessary withdrawal of disease-modifying therapies^18,19^.

Finally, regarding patient access to potassium monitoring as recommended by international clinical guidelines, a substantial perceived public–private gap was identified (55.5% vs 90.3%), predominantly reported by physicians practicing in Guatemala, El Salvador, Panama, Honduras, and Nicaragua. This observation aligns with socioeconomic reports from Latin America highlighting limited access to high-quality healthcare across multiple dimensions, including availability, affordability, physical access, and acceptability^20^. This represents a particularly relevant finding of this survey, as it has direct implications for engagement with healthcare decision-makers in public health systems across the region. These findings suggest opportunities for organizational and system-level improvements to ensure compliance with recommended 24- hour reassessment (potassium 6.0–6.4 mEq/L) and immediate action with ECG monitoring (potassium ≥6.5 mEq/L) as outlined in international and regional guidance ^6,7^.

Study limitations include restricting participation to physicians in the sponsor’s database across Central America and the Dominican Republic, which may limit generalizability to all specialists and bias towards answers selected. Voluntary, electronic invitations may underrepresent physicians in resource-limited or remote settings. Limited participation from countries like Nicaragua and El Salvador further constrains country-specific conclusions. Nevertheless, given the diversity of specialties, lack of specialty-by-country disaggregation, and relatively homogeneous context across the region, these limitations are unlikely to substantially affect the overall findings.

In conclusion, this study provides insight into current attitudes and practices related to the diagnosis and management of HK in Central America and the Dominican Republic, highlighting areas of opportunity in emergency referral criteria, RAASi reduction and access to serum potassium monitoring. Overall, the results demonstrate partial alignment with consensus statements and guidelines, alongside identifiable and potentially addressable gaps in clinical practice.

## Data Availability

Datasets related to the study are available to a reasonable request and can be solicited at

## Acknowledgments

The authors would like to thank all the physicians who responded to the survey for their time and expertise; Dr. Roy Wong and Ms. Diana Paniagua, representatives of Data Analytic, for their support during the execution of this project; and Drs. Andres Lancini and Marcelle Elias for their assistance in the drafting and review of this manuscript.

## Author Contributions

Study conceptualization and design: Avellán M, González P. Instrument development and data collection: Ortíz D, González J. Data analysis: Avellán M, Sánchez-Polo V. Drafting of the introduction and discussion: Avellán M; González J. Critical revision of the intellectual content: All authors. Study supervision: Dr. Marta Avellán.

## Ethical Considerations

This study was sponsored by AstraZeneca CAMCAR and received internal institutional approval. Physician participation was voluntary, with electronic informed consent included at the beginning of the questionnaire. No identifiable data was collected, and no financial incentives were offered. Confidentiality of responses was ensured at all times.

## —SUPLEMENTARY MATERIAL—

## Appendix 1. Informed Consent – English Version

### Study Title

Survey on Perceptions and Therapeutic Attitudes Toward the Management of Hyperkalemia/Hyperpotassemia in Adults by Medical Specialists Practicing in Central America and the Dominican Republic during the period April–June 2025.

### Principal Investigators

Dr. Marta Avellán, Dr. Pablo González, Dr. Dean Ortiz, Dr. Josué González, Dr. Vicente Sánchez- Polo.

### Study Sponsor

This study is being conducted by the company Data Analytic and sponsored by AstraZeneca.

### Purpose of the Study

The purpose of this study is to better understand how medical specialists in Central America and the Dominican Republic perceive and manage hyperkalemia/hyperpotassemia in adult patients. This study will be conducted between April and June 2025.

### Study Procedures

You are invited to participate in a study consisting of completing an online questionnaire with 16 questions. These questions have been carefully validated by experts and peer-reviewed to ensure their relevance and accuracy.

### Duration of Participation

Completion of the questionnaire will take approximately 15 to 20 minutes.

### Confidentiality

We are committed to ensuring that your participation in this study is completely confidential. All responses you provide will be collected anonymously and aggregated for analytical purposes. At no time will your name be disclosed or your individual responses shared with third parties. All information will be used exclusively for research purposes and in compliance with applicable data protection regulations.

### Voluntary Participation

Your participation in this study is entirely voluntary, and you may withdraw at any time without any consequences. No financial compensation will be provided for participation in this study. However, your contribution will be invaluable in improving medical practices related to the management of hyperkalemia in Central America and the Dominican Republic.

### Risks and Benefits

No significant risks are anticipated from participating in this study. Although you will not receive direct benefits, your participation will contribute to improving knowledge regarding the management of hyperkalemia/hyperpotassemia in the region.

### Contact for Questions

If you have any questions regarding the study, you may contact us at

### Statement of Consent

I have read and understood the information provided and agree to participate in the study.* Select only one answer:

- Yes
- No

## Appendix 2. Survey – English Version

### I. Respondent Identification Data

Please select the medical specialty you primarily practice as a healthcare professional* Select one of the following options. Select only one answer.

- Cardiology
- Nephrology
- Internal Medicine
- Endocrinology
- Geriatrics
- General Medicine
- Other

Please select the country where you primarily practice* Select one of the following options. Select only one answer.

- Guatemala
- Honduras
- Nicaragua
- El Salvador
- Costa Rica
- Panama
- Dominican Republic

### II General Questions on Physicians’ Perception and Attitudes Toward Hyperkalemia

In your opinion, at what serum potassium concentration would you consider the diagnosis of hyperkalemia?*

Select one of the following options. Select only one answer.

- ≥5.0 mEq/L
- ≥5.5 mEq/L
- ≥6.0 mEq/L
- ≥6.5 mEq/L
- ≥7.0 mEq/L

In your opinion, at what serum potassium concentration does hyperkalemia warrant outpatient medical intervention (e.g., modification of current treatment, dietary changes, use of loop diuretics or ion-exchange resins, referral to another specialist, etc.)?* Select one of the following options. Select only one answer.

- ≥5.0 mEq/L
- ≥5.5 mEq/L
- ≥6.0 mEq/L
- ≥6.5 mEq/L
- ≥7.0 mEq/L
- Upon evidence of clinical or electrocardiographic changes associated with elevated serum potassium

In your opinion, at what serum potassium concentration would you consider referring your patient to the emergency department for management of hyperkalemia?* Select one of the following options. Select only one answer.

- ≥5.0 mEq/L
- ≥5.5 mEq/L
- ≥6.0 mEq/L
- ≥6.5 mEq/L
- ≥7.0 mEq/L
- Upon evidence of clinical or electrocardiographic changes associated with elevated serum potassium

Is it routine in your practice to perform an electrocardiogram before deciding whether your patient requires an intervention for elevated serum potassium?* Select only one answer.

- Yes
- No

Is it routine in your practice to recommend avoidance of foods with high potassium content as treatment for hyperkalemia?* Select only one answer.

- Yes
- No

Is it routine in your practice to reduce or withhold medications that may be increasing serum potassium when your patient develops hyperkalemia?* Select one of the following options. Select only one answer.

- Yes, I discontinue the medication
- Yes, I reduce the dose without discontinuing
- No

Is it routine in your practice to prescribe medications that increase potassium excretion (e.g., loop diuretics) during the management of hyperkalemia?* Select only one answer.

- Yes
- No

Based on your clinical experience, please select the frequency level of the following causes of hyperkalemia* Please select the appropriate response for each item:

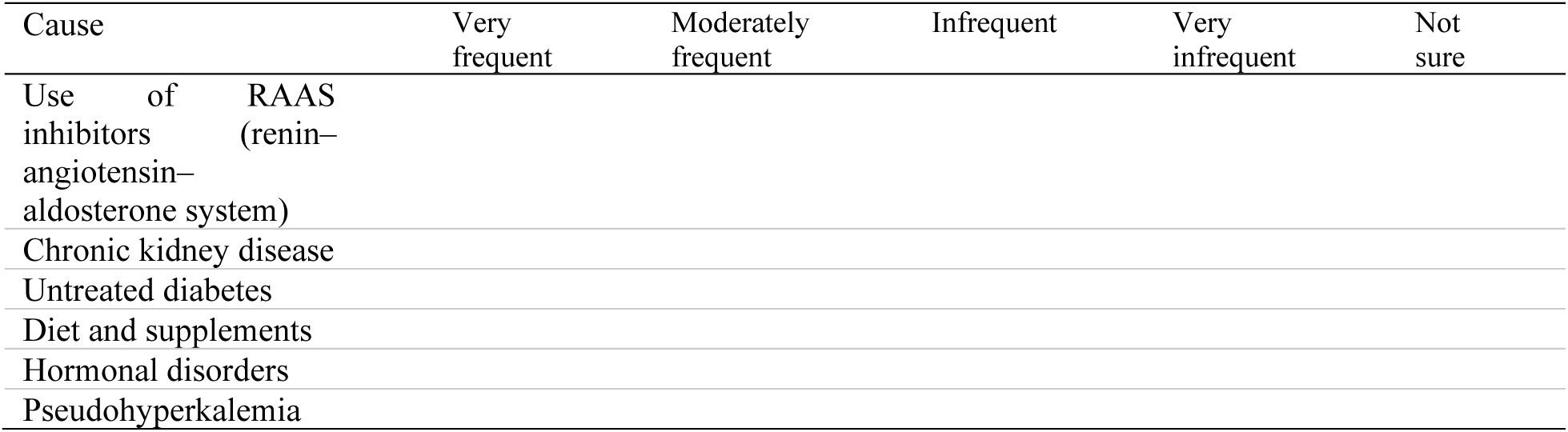

Do you consider that ALL of your patients have access to potassium monitoring as recommended by clinical practice guidelines?* Please select the appropriate response for each setting:

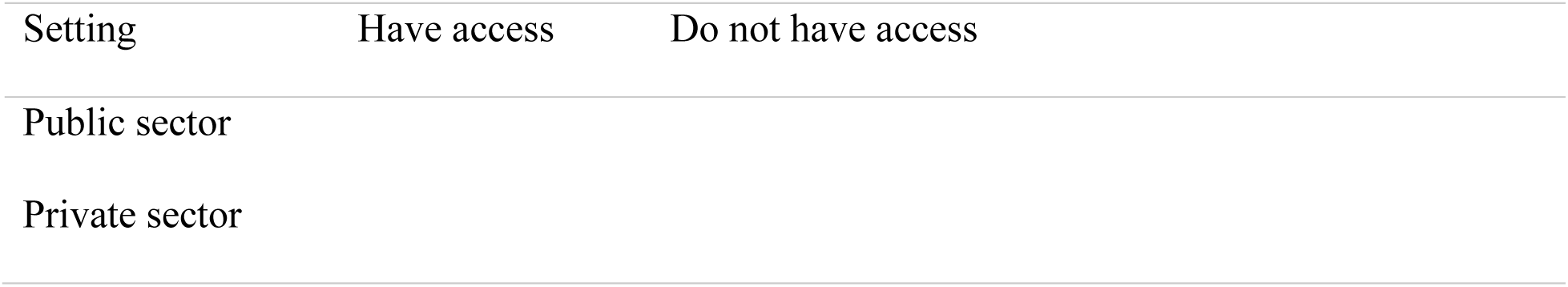

Please respond according to the public or private healthcare setting, as applicable. According to your clinical experience, how frequent is hyperkalemia among the patients you routinely manage?* Select one of the following options. Select only one answer.

- Less than 10%
- Between 10% and 30%
- Between 30% and 50%
- More than 50%
- I am not sure of the percentage

### III Perception Toward Hyperkalemia Among Cardiologists in Patients With Heart Failure

Based on your clinical experience, how frequent is hyperkalemia in your patients with heart failure?* Select one of the following options. Select only one answer.

- Less than 10%
- Between 10% and 30%
- Between 30% and 50%
- More than 50%
- I am not sure of the percentage

Currently, which pharmacological strategies do you use most frequently to manage elevated serum potassium in patients with heart failure without electrocardiographic changes?

### Multiple choice

Please select the appropriate response for each item:

| Strategy | Use frequently | Use occasionally | Do not use |
| --- | --- | --- | --- |
| Reduce/withhold mineralocorticoid receptor antagonists |  |  |  |
| Reduce/withhold ACE inhibitors/ARBs |  |  |  |
| Reduce/withhold neprilysin inhibitors |  |  |  |
| Initiate loop diuretics |  |  |  |
| Use of cation-exchange resins |  |  |  |
| Other (specify) |  |  |  |
| Other (any strategy not listed above), please specify: [Open text] |  |  |  |

If you were to rate your level of concern regarding hyperkalemia in your patients with heart failure, which of the following options would you choose?*

- A major concern
- A secondary or minor concern
- Not a concern
- Perception Toward Hyperkalemia Among Nephrologists in Patients With Chronic Kidney Disease

Based on your clinical experience, how frequent is hyperkalemia in your patients with stage 4–5 chronic kidney disease who are not yet receiving renal replacement therapy?* Select one of the following options. Select only one answer.

- Less than 10%
- Between 10% and 30%
- Between 30% and 50%
- More than 50%
- I am not sure of the proportion

Currently, which strategies do you use most frequently to manage elevated serum potassium in patients with stage 4–5 chronic kidney disease who do not yet require renal replacement therapy?

### Multiple choice

Please select the appropriate response for each item:

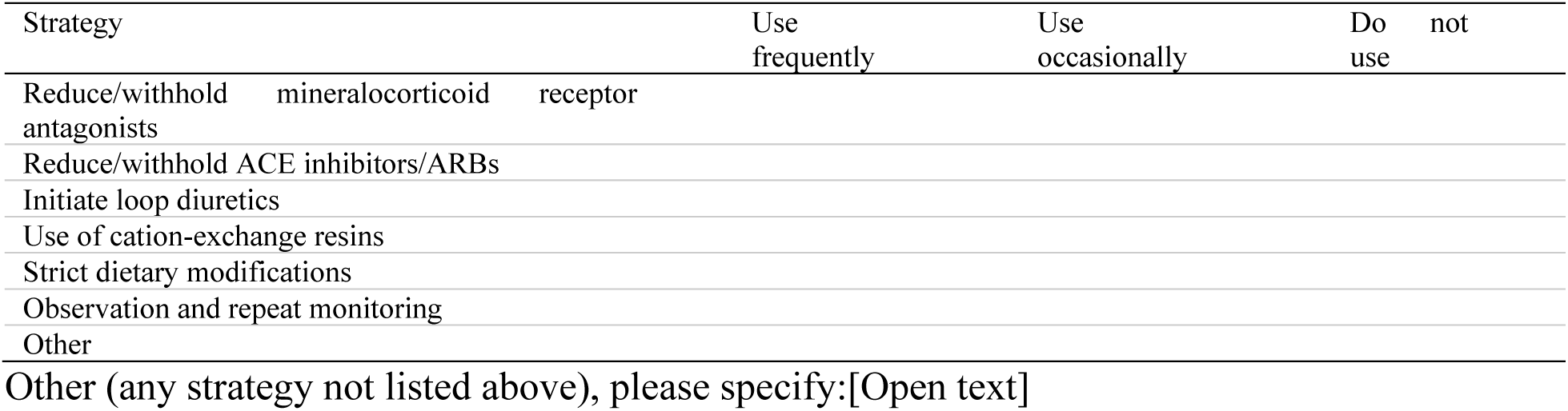

Thank you very much for participating in the study on perception and therapeutic attitudes toward the management of hyperkalemia. Your participation is very important to us!

---

**Supplementary Table 1.**
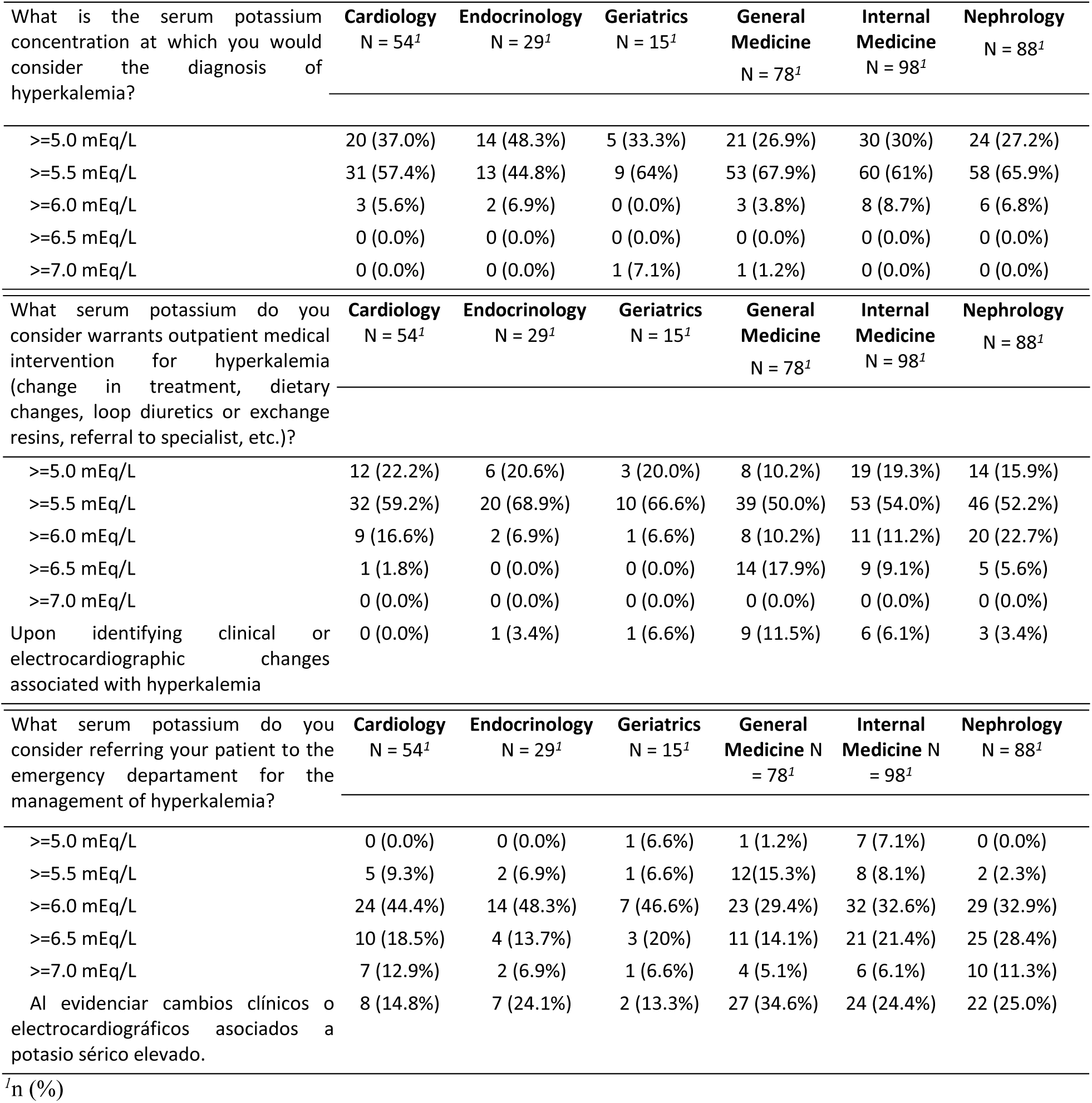
Distribution of Surveyed Physicians According to potassium thresholds for the diagnosis of hyperkalemia.

**Supplementary Table 2.** Distribution of Perceived Causes of Hyperkalemia Among Physicians by Specialty.

| Causes of Hyperkalemia | Overall N = 362<sup>†</sup> | Cardiology N = 54<sup>†</sup> | Endocrinology N = 29<sup>†</sup> | Geriatrics N = 15<sup>†</sup> | General Medicine N = 78<sup>†</sup> | Internal Medicine N = 98<sup>†</sup> | Nephrology N = 88<sup>†</sup> |
| --- | --- | --- | --- | --- | --- | --- | --- |
| **Use of RAAS Inhibitors** |  |  |  |  |  |  |  |
| Moderately frequent | 133 (36.7%) | 23 (42.6%) | 9 (31.0%) | 5 (33.3%) | 31 (39.7%) | 37 (37.8%) | 28 (31.8%) |
| Very frequent | 63 (17.4%) | 15 (27.8%) | 3 (10.3%) | 4 (26.7%) | 7 (9.0%) | 16 (16.3%) | 18 (20.5%) |
| Very infrequent | 46 (12.7%) | 3 (5.6%) | 6 (20.7%) | 1 (6.7%) | 11 (14.1%) | 13 (13.3%) | 12 (13.6%) |
| Not sure | 6 (1.7%) | 0 (0.0%) | 0 (0.0%) | 0 (0.0%) | 3 (3.8%) | 1 (1.0%) | 2 (2.3%) |
| Infrequent | 114 (31.5%) | 13 (24.1%) | 11 (37.9%) | 5 (33.3%) | 26 (33.3%) | 31 (31.6%) | 28 (31.8%) |
| **Chronic Kidney Disease** |  |  |  |  |  |  |  |
| Moderately frequent | 86 (23.8%) | 12 (22.2%) | 11 (37.9%) | 7 (46.7%) | 20 (25.6%) | 13 (13.3%) | 23 (26.1%) |
| Very frequent | 263 (72.7%) | 41 (75.9%) | 15 (51.7%) | 8 (53.3%) | 50 (64.1%) | 85 (86.7%) | 64 (72.7%) |
| Very infrequent | 3 (0.8%) | 0 (0.0%) | 0 (0.0%) | 0 (0.0%) | 3 (3.8%) | 0 (0.0%) | 0 (0.0%) |
| Not sure | 3 (0.8%) | 0 (0.0%) | 0 (0.0%) | 0 (0.0%) | 3 (3.8%) | 0 (0.0%) | 0 (0.0%) |
| Infrequent | 7 (1.9%) | 1 (1.9%) | 3 (10.3%) | 0 (0.0%) | 2 (2.6%) | 0 (0.0%) | 1 (1.1%) |
| **Untreated Diabetes** |  |  |  |  |  |  |  |
| Moderately frequent | 113 (31.2%) | 18 (33.3%) | 3 (10.3%) | 8 (53.3%) | 28 (35.9%) | 26 (26.5%) | 30 (34.1%) |
| Very frequent | 49 (13.5%) | 5 (9.3%) | 2 (6.9%) | 2 (13.3%) | 24 (30.8%) | 5 (5.1%) | 11 (12.5%) |
| Very infrequent | 70 (19.3%) | 7 (13.0%) | 16 (55.2%) | 2 (13.3%) | 6 (7.7%) | 27 (27.6%) | 12 (13.6%) |
| Not sure | 7 (1.9%) | 2 (3.7%) | 0 (0.0%) | 0 (0.0%) | 4 (5.1%) | 1 (1.0%) | 0 (0.0%) |
| Infrequent | 123 (34.0%) | 22 (40.7%) | 8 (27.6%) | 3 (20.0%) | 16 (20.5%) | 39 (39.8%) | 35 (39.8%) |
| **Diet and Supplements** |  |  |  |  |  |  |  |
| Moderately frequent | 107 (29.6%) | 19 (35.2%) | 5 (17.2%) | 4 (26.7%) | 20 (25.6%) | 18 (18.4%) | 41 (46.6%) |
| Very frequent | 47 (13.0%) | 5 (9.3%) | 0 (0.0%) | 0 (0.0%) | 16 (20.5%) | 12 (12.2%) | 14 (15.9%) |
| Very infrequent | 58 (16.0%) | 3 (5.6%) | 14 (48.3%) | 3 (20.0%) | 6 (7.7%) | 22 (22.4%) | 10 (11.4%) |
| Not sure | 9 (2.5%) | 2 (3.7%) | 0 (0.0%) | 1 (6.7%) | 6 (7.7%) | 0 (0.0%) | 0 (0.0%) |
| Infrequent | 141 (39.0%) | 25 (46.3%) | 10 (34.5%) | 7 (46.7%) | 30 (38.5%) | 46 (46.9%) | 23 (26.1%) |
| **Hormonal Disorders** |  |  |  |  |  |  |  |
| Moderately frequent | 89 (24.6%) | 9 (16.7%) | 9 (31.0%) | 3 (20.0%) | 17 (21.8%) | 30 (30.6%) | 21 (23.9%) |
| Very frequent | 23 (6.4%) | 3 (5.6%) | 4 (13.8%) | 1 (6.7%) | 7 (9.0%) | 4 (4.1%) | 4 (4.5%) |
| Very infrequent | 70 (19.3%) | 10 (18.5%) | 4 (13.8%) | 2 (13.3%) | 11 (14.1%) | 23 (23.5%) | 20 (22.7%) |
| Not sure | 19 (5.2%) | 4 (7.4%) | 0 (0.0%) | 0 (0.0%) | 7 (9.0%) | 4 (4.1%) | 4 (4.5%) |
| Infrequent | 161 (44.5%) | 28 (51.9%) | 12 (41.4%) | 9 (60.0%) | 36 (46.2%) | 37 (37.8%) | 39 (44.3%) |
| **Pseudohyperkalemia** |  |  |  |  |  |  |  |
| Moderately frequent | 100 (27.6%) | 14 (25.9%) | 10 (34.5%) | 3 (20.0%) | 24 (30.8%) | 32 (32.7%) | 17 (19.3%) |
| Very frequent | 29 (8.0%) | 2 (3.7%) | 1 (3.4%) | 1 (6.7%) | 5 (6.4%) | 8 (8.2%) | 12 (13.6%) |
| Very infrequent | 72 (19.9%) | 10 (18.5%) | 7 (24.1%) | 2 (13.3%) | 12 (15.4%) | 17 (17.3%) | 24 (27.3%) |
| Not sure | 36 (9.9%) | 10 (18.5%) | 2 (6.9%) | 3 (20.0%) | 10 (12.8%) | 10 (10.2%) | 1 (1.1%) |
| Infrequent | 125 (34.5%) | 18 (33.3%) | 9 (31.0%) | 6 (40.0%) | 27 (34.6%) | 31 (31.6%) | 34 (38.6%) |

<sup>†</sup>n (%)

**Supplementary Table 3.** Distribution of Surveyed Physicians According to Initial Management Strategies for Hyperkalemia.

| Hyperkalemia Management | Overall<br>N = 362<sup>1</sup> | Cardiology<br>N = 54<sup>1</sup> | Endocrinology<br>N = 29<sup>1</sup> | Geriatrics<br>N = 15<sup>1</sup> | General<br>Medicine N<br>= 78<sup>1</sup> | Internal<br>Medicine N<br>= 98<sup>1</sup> | Nephrology<br>N = 88<sup>1</sup> |
| --- | --- | --- | --- | --- | --- | --- | --- |
| **Requires an electrocardiogram before deciding on intervention for elevated serum potassium** |  |  |  |  |  |  |  |
| No | 121 (33.4%) | 15 (28%) | 13 (45%) | 10 (67%) | 21 (27%) | 27 (28%) | 35 (40%) |
| Yes | 241 (67.0%) | 39 (72%) | 16 (55%) | 5 (33.3%) | 57 (73%) | 71 (72%) | 53 (60%) |
| **Reduce dose or discontinue medications that may be increasing serum potassium** |  |  |  |  |  |  |  |
| No | 26 (7.2%) | 2 (3.7%) | 2 (6.9%) | 0 (0.0%) | 6 (7.7%) | 6 (6.1%) | 10 (11.3%) |
| Yes, I discontinue the medication | 143 (39.5%) | 23 (42.6%) | 9 (31.0%) | 4 (27.0%) | 24 (31.0%) | 56 (57.1%) | 27 (31.0%) |
| Yes, I reduce the dose without discontinuing | 193 (53.3%) | 29 (54%) | 18 (62.1%) | 11 (73.3%) | 48 (61.5%) | 36 (37.0%) | 51 (58.0%) |
| **Prescription of medications that increase potassium excretion** |  |  |  |  |  |  |  |
| No | 95 (26.2%) | 10 (18.5%) | 11 (37.9%) | 3 (20.0%) | 34 (43.6%) | 22 (22.4%) | 15 (17.0%) |
| Yes | 267 (74.0%) | 44 (81.5%) | 18 (62.0%) | 12 (80.0%) | 44 (56.4%) | 76 (77.5%) | 73 (83.0%) |

<sup>1</sup>n (%)

**Supplementary Table 4.** Accessibility to Potassium Monitoring in the Follow-up of Clinical Guideline Recommendations by Healthcare Sector.

| Accessibility | Overall<br>N = 362<sup>1</sup> | Costa<br>Rica N =<br>117<sup>1</sup> | El<br>Salvador<br>N = 13<sup>1</sup> | Guatemala<br>N = 76<sup>1</sup> | Honduras<br>N = 101<sup>1</sup> | Nicaragua<br>N = 7<sup>1</sup> | Panamá<br>N = 23<sup>1</sup> | Dominican<br>Republic N =<br>25<sup>1</sup> |
| --- | --- | --- | --- | --- | --- | --- | --- | --- |
| **Public Sector** |  |  |  |  |  |  |  |  |
| Do not have<br>accessibility | 161<br>(44.5%) | 31<br>(26.5%) | 7 (53.8%) | 37 (48.7%) | 62 (61.4%) | 5 (71.4%) | 12<br>(52.2%) | 7 (28.0%) |
| Have<br>accessibility | 201<br>(55.5%) | 86<br>(73.5%) | 6 (46.2%) | 39 (51.3%) | 39 (38.6%) | 2 (28.6%) | 11<br>(47.8%) | 18 (72.0%) |
| **Private Sector** |  |  |  |  |  |  |  |  |
| Do not have<br>accessibility | 35<br>(9.7%) | 15<br>(12.8%) | 0 (0.0%) | 12 (15.8%) | 8 (7.9%) | 0 (0.0%) | 0 (0.0%) | 0 (0.0%) |
| Have<br>accessibility | 327<br>(90.3%) | 102<br>(87.2%) | 13<br>(100.0%) | 64 (84.2%) | 93 (92.1%) | 7 (100.0%) | 23<br>(100.0%) | 25 (100.0%) |

<sup>1</sup>n (%)

|  | Cardiology<br>N = 54<sup>1</sup> | Endocrinology<br>N = 29<sup>1</sup> | Geriatrics<br>N = 15<sup>1</sup> | General<br>Medicine<br>N = 78<sup>1</sup> | Internal<br>Medicine<br>N = 98<sup>1</sup> | Nephrology<br>N = 88<sup>1</sup> |
| --- | --- | --- | --- | --- | --- | --- |
| **Public Sector** |  |  |  |  |  |  |
| Do not have<br>accessibility | 20 (37.0%) | 8 (28.0%) | 6 (40.0%) | 45<br>(57.7%) | 48 (49.0%) | 35 (40.0%) |
| Have<br>accessibility | 34 (63.0%) | 21 (72.4%) | 9 (60.0%) | 33<br>(42.3%) | 50 (51.0%) | 53 (60.2%) |
| **Private Sector** |  |  |  |  |  |  |
| Do not have<br>accessibility | 2 (3.7%) | 2 (6.9%) | 3 (20.0%) | 10<br>(12.8%) | 5 (5.4%) | 10 (11.3%) |
| Have<br>accessibility | 52 (96.3%) | 27 (93.1%) | 12 (80.0%) | 68<br>(87.2%) | 93 (94.9%) | 78 (88.6%) |

<sup>1</sup>n (%)

**Supplementary Table 5.**
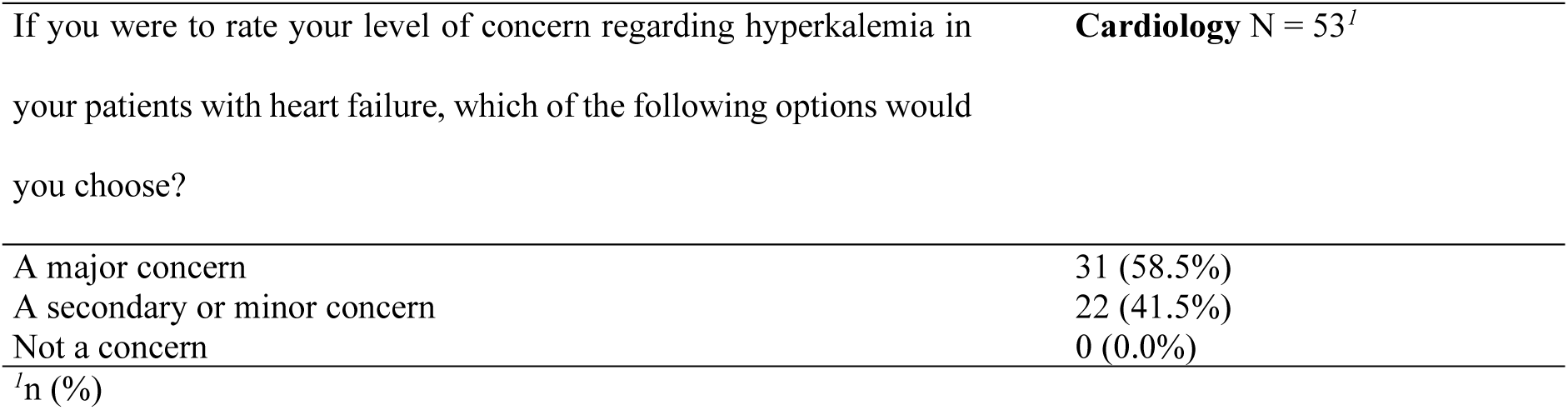
Distribution of Perceived Concern regarding Hyperkalemia among cardiologist.

